# Health service delivery gaps and support systems for addressing central obesity in women beyond the postpartum period

**DOI:** 10.64898/2026.02.05.26345697

**Authors:** Rosela Remigius, Rosula Remigius, Zalia O. Basheikh

## Abstract

**Background:** Central obesity is a critical public health issue linked to non- communicable diseases and long-term maternal health risks. Women beyond the postpartum period often face barriers to weight management, yet limited evidence exists on their specific challenges and the role of healthcare systems in addressing them. This study aimed to assess central obesity among women beyond postpartum period, focusing on the associated challenges and available health support systems.

**Methodology:** A cross-sectional design was employed whereby 120 women (1 to 5 years postpartum), aged 18 - 49 years, attending selected one private and two government health facilities in Morogoro Urban District participated. Face-to-face interviews using semi-structured questionnaires and anthropometric measurements of waist and hip circumference were conducted. Data were recorded using Kobo digital Toolbox.

**Key findings:** High prevalence of central obesity, with 68.3% of participants having a waist circumference above normal (≥80 cm) was observed. Although no socio-demographic factor assessed showed a statistically significant association with central obesity, trends indicated higher odds among older (OR=1.544; 95% CI, 0.084-28.557), married (OR= 1.730; 95% CI, 0.612-4.892) and higher income women (OR= 4.878; 95% CI, 0.367-64.818). Lifestyle behaviors such as low physical activity, poor dietary habits and lack of portion control were prevalent. 57.5% reported lacking information on weight and waist management. Despite attending health care facilities, 94.2% of the women had never received guidance from health providers regarding weight or central obesity management and 95% reported not receiving any form of support such as nutrition counseling or exercise recommendations.

**Conclusion:** The study concludes that central obesity is highly prevalent among women beyond postpartum period and is influenced by poor lifestyle behaviors and inadequate healthcare system support. It recommends integrating weight management strategies into routine postpartum care and strengthening healthcare systems to offer tailored guidance and support to women after childbirth.

## Introduction

Obesity and non-communicable diseases (NCDs) are closely linked (1). The NCDs including cancer, stroke, heart disease, diabetes and chronic lung disease, are the major disease burden worldwide that collectively contribute to 71% of mortality globally (2). According to Wang et al. (3) they reported that, with the economic development, prevalent sedentary lifestyle, dietary transition and increasing aging population worldwide, it is expected that the obesity and NCDs burden will continue to rise and so affect people of all ages from all social groups in both developed and developing countries.

Obesity as defined by the World Health Organization (WHO) (4), is an abnormal or excess fat accumulation that may impair health. It is associated with the imbalance between intake and expenditure of energy (5). This caloric imbalance creates an excess accumulation of energy that in turn is stored and resulting to excess body weight (6). Fat distribution plays a crucial role in determining metabolic risk and based on the location of fat accumulation, there are two types of body forms which obese individuals possess and these are: fat accumulation in lower body such as hips and thighs commonly known as pear shape or gynecoid and fat accumulation in upper body such as the abdominal region commonly known as apple-shaped or android (7). However, abdominal obesity also known as central obesity is considered as the more serious form of fat distribution among the obese individuals because it poses individuals to various metabolic disorders and diseases (8).

The common anthropometric measurements used to assess central obesity are; Waist circumference (WC), Waist-to-hip ratio (WHR) and Waist-to-height-ratio (WHTR). Most studies have used WC as the defining criterion because it is convenient and strongly related with intraabdominal fat content and cardiovascular risk factors (9,10).. Basing on WC, the cutoff points differ depending on various definitions provided by the World Health Organization (WHO), International Diabetes Federation (IDF) and the Adult Treatment Panel III guidelines (ATP III) (11). According to (12), the cutoff points definitions vary according to sex, for example among men, there are three cutoff points which are ≥ 90 cm or ≥ 94 cm or ≥ 102 cm and for women there are two which are ≥ 80 cm and ≥ 88 cm. But also, the threshold of waist circumference used to define central obesity depends on ethnic groups and the worlds regions where, for Sub-Saharan Africa WHO defined abdominal obesity by fixing specific waist circumference cutoff points at ≥ 94 cm for men and ≥ 80 cm for women.

Central obesity contributes to an increased risk of insulin resistance, type 2 diabetes, metabolic syndrome, cardiovascular diseases, chronic respiratory diseases and it causes mortality (13,14). It also results to various types of cancer example colorectum, pancreas, endometrium and breast cancer (15). Central obesity is also associated with other co-morbidities such as dyslipidemia, hip fracture and depression (16).

Women have been observed to be at a higher risk of central obesity compared to other groups as globally, the prevalence of central obesity is estimated to be higher among females (47.6%) than males (30.4%). This is due to the gender difference whereby females generally have higher body fat proportion than males (17). Studies made in Uganda (18), Nigeria (19) and Cote d’lvoire (20) reported that women had higher prevalence rate of central obesity than men. And a study conducted in South Africa reported that one in three adults of normal weight had central obesity (21). In Tanzania mainland, according to Ministry of health in 2022, the prevalence of overweight or obesity among women was 36%. Among those aged 20 to 49 years, overweight and obesity increased with education and household wealth. Also, for central obesity a study conducted in Dodoma city, found high prevalence of central obesity among females (67.4%) compared to males (28.0%) (22).

Most women beyond postpartum period have been reported to face barriers in management of weight which include limited time due to infant care responsibilities, lack of support from partners and being less prioritized where their needs come the last in family (23). According to (24) high uncontrolled eating is linked with central obesity in women after pregnancy and most maternal health guidelines around the world do not address postpartum weight or lifestyle (25) and so there is a need for clear implementation strategies to guide evidence translation into practice to deliver public health impact (26).

However, there is limited data and information on the barriers that women beyond postpartum period face and the adequacy of healthcare systems in managing central obesity. This knowledge gap hinders the development of effective prevention and management strategies of central obesity among these women. Thus, it is crucial to investigate the challenges and support systems available for women beyond the post-partum period to inform the targeted interventions. This study aimed to assess factors influencing and challenges towards management of central obesity among women beyond postpartum as well as the role of health care systems in prevention and management.

## Materials and methodology

### Description of study site

This study was carried out in Morogoro Urban District in Morogoro Region, Tanzania. According to the 2022 population census, Morogoro Urban District has a population of 471,409 (City Population, 2023). (27). Administratively, the district is divided into 29 wards, including Bigwa, Boma, Chamwino, Kauzeni, Kichangani, Kihonda, Kihonda Magorofani, Kilakala, Kingo, Kingolwira, Kiwanja cha Ndege, Luhungo, Lukobe, Mafiga, Mafisa, Magadu, Mazimbu, Mbuyuni, Mindu, Mji Mkuu, Mji Mpya, Mkundi, Mlimani, Mwembesongo, Mzinga, Sabasaba, Sultan Area, Tungi, and Uwanja wa Taifa.

Health services in Morogoro Region are provided through public and private institutions like the regional referral hospital, district hospitals, health centers, dispensaries, faith-based hospitals and private hospitals. In total, the region has approximately 419 operational health facilities, of which 354 are dispensaries, 52 are health centers and 13 are hospitals. These facilities offer a variety of services, including outpatient care, surgery and diagnostic services such as X-ray, ultrasound, and laboratory tests (27).

### Study design and sampling procedure

#### Study design

A cross-sectional design was used, where data were collected only once from the selected women beyond postpartum attending the selected health facilities from 15^th^ April 2025 to 13^th^ June 2025.

#### Study population

This study focused on women beyond postpartum period who were between 1 to 5 years postpartum. The inclusion criteria for the study population were women aged 18 to 49 years (childbearing age) who had delivered a live birth. The exclusion criteria were women who had experienced stillbirth or miscarriage, as their experiences might have differed significantly and women who were pregnant again during the study period.

#### Sampling design

The study employed non-probability sampling techniques. Morogoro Urban District was purposely selected due to its logistical feasibility within the three-month study period and its suitability for capturing data from an urban population, which aligns with the research focus.. Three health facilities Morogoro Polyclinic, Uhuru and SabaSaba were selected. They were selected because of offering a wide range of health services, specifically maternal and child health services, which facilitated the collection of relevant information and provided insights into the support systems available for postpartum women.

Using convenient sampling, eligible women beyond postpartum visiting the selected health facilities during the study period were approached and invited to participate. Also, the key informants who were healthcare providers (nurses, nutritionists and doctors) were purposely selected based on their professional expertise and experience in providing care to postpartum women.

A total of 192 women beyond postpartum were involved in the study, representing half of the sample size obtained from the Cochran’s formula (Equation. 1). The adjustment was necessary due to limited time and resources.

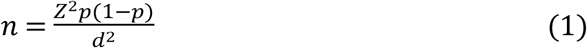

Where;

n = desired sample size

z = standard normal deviation set at 1.96 corresponding to 95% confidence interval

p = proportion of women beyond postpartum with central obesity used here is 50% (0.5) because there is no prior data on actual proportion of women beyond postpartum affected by central obesity.

q = (1-p) proportion of women beyond postpartum who are estimated to not have central obesity which is 0.5

d = degree of accuracy desired (0.05)

Therefore;

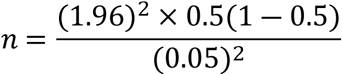

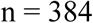

The sample size of women beyond postpartum 354 is then divided by 2

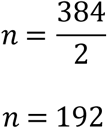

Therefore; the sample size of women beyond postpartum was 192.

To ensure that each health facility was adequately represented in the study, the sample size was proportionally allocated based on the number of eligible women beyond postpartum attending each health facility. Data were obtained from health facility records on the average monthly number of women beyond postpartum visiting each facility. The total number of eligible women across all selected health facilities was then calculated. Thereafter, the proportion of the total sample size assigned to each facility was determined by dividing the number of eligible women at that specific facility by the total number of eligible women across all facilities, using the formula below:

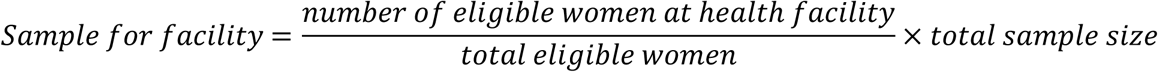

### Data collection

Primary quantitative and qualitative data were collected from women beyond postpartum and key informants (nurses, nutritionists and doctors) through in-depth face-to-face interviews, guided by two semi-structured questionnaires. Data collection was conducted using Kobo Toolbox, a digital platform for mobile data collection. The first questionnaire was used for women beyond postpartum and it focused on demographic information, lifestyle, health care experiences, self-perceptions, knowledge and challenges faced by women beyond postpartum. The second questionnaire was used for the key informant interviews. Anthropometric measures such as waist and hip circumference were also taken. The measurements were taken using a flexible, non-stretchable measuring tape. Participants stood upright with their arms at their sides, and the tape was placed around the narrowest part of the abdomen for waist circumference and around the widest part for hip circumference. Waist circumference was recorded in centimeters, and according to WHO (2021) cut-off points, a measurement of **≥** 80 cm was classified as central obesity. For the WHR a measurement of ≥ 0.85 was classified as central obesity.

### Data analysis

Data were entered, managed and analyzed using the Statistical Package for Social Sciences (SPSS) version 21 and Microsoft Excel. Descriptive statistics including means, frequencies and percentages were calculated. Frequencies and percentages were used to describe categorical variables such as participants’ age groups, the proportion of participants with central obesity, parity and health facility distribution. Means were used to summarize continuous variables such as the age of participants, waist circumference and the hip circumference of the women beyond postpartum. Inferential statistics were employed to determine the association between socio-demographic characteristics (marital status, education level, parity, occupation, age, and income) and central obesity using logistic regression analysis.

### Ethical consideration

This study involved human participants and was conducted in accordance with the ethical principles of the Declaration of Helsinki. Ethical approval for the study was obtained from the relevant institutional ethics review committee prior to data collection Sokoine University of Agriculture, with approval number SUA/DOS/R.1/1. Permission to conduct the study was obtained from the respective health facilities in Morogoro Urban District. All eligible participants were provided with clear information about the purpose, procedures, potential benefits and minimal risks of the study. Participation was entirely voluntary, and participants were informed of their right to decline or withdraw from the study at any time without any consequences to the health services they receive. Verbal informed consent was obtained from all participants before participation and those who accepted where the ones whose data were documented and taken from the field. The use of verbal consent was approved by the ethics committee, as the study involved minimal risk and did not include invasive procedures. Consent was documented by the research team prior to data collection. Participants’ privacy and confidentiality were strictly maintained. No personal identifiers were collected, and all data were anonymized and securely stored with access restricted to the research team.

## Results

### Socio-demographic characteristics

More than half (55%) of mothers beyond postpartum were aged 20-29 years. Majority (80.8%) were married or living with the partner, and had 1 to 2 children (74%). Also, nearly half of subjects (44.2%) had secondary level of education, 62% were self-employed doing small business and 49.2% had medium monthly income (Table 1).

**Table 1.**
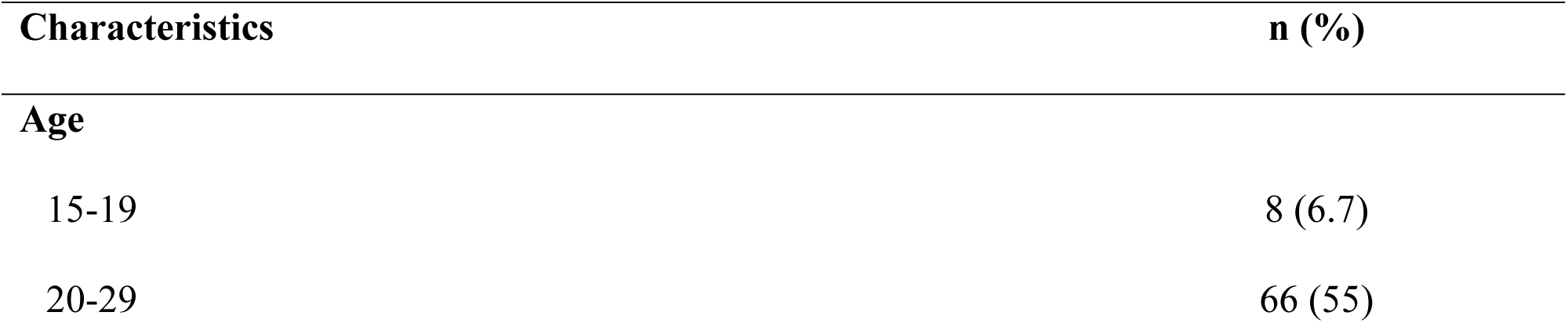
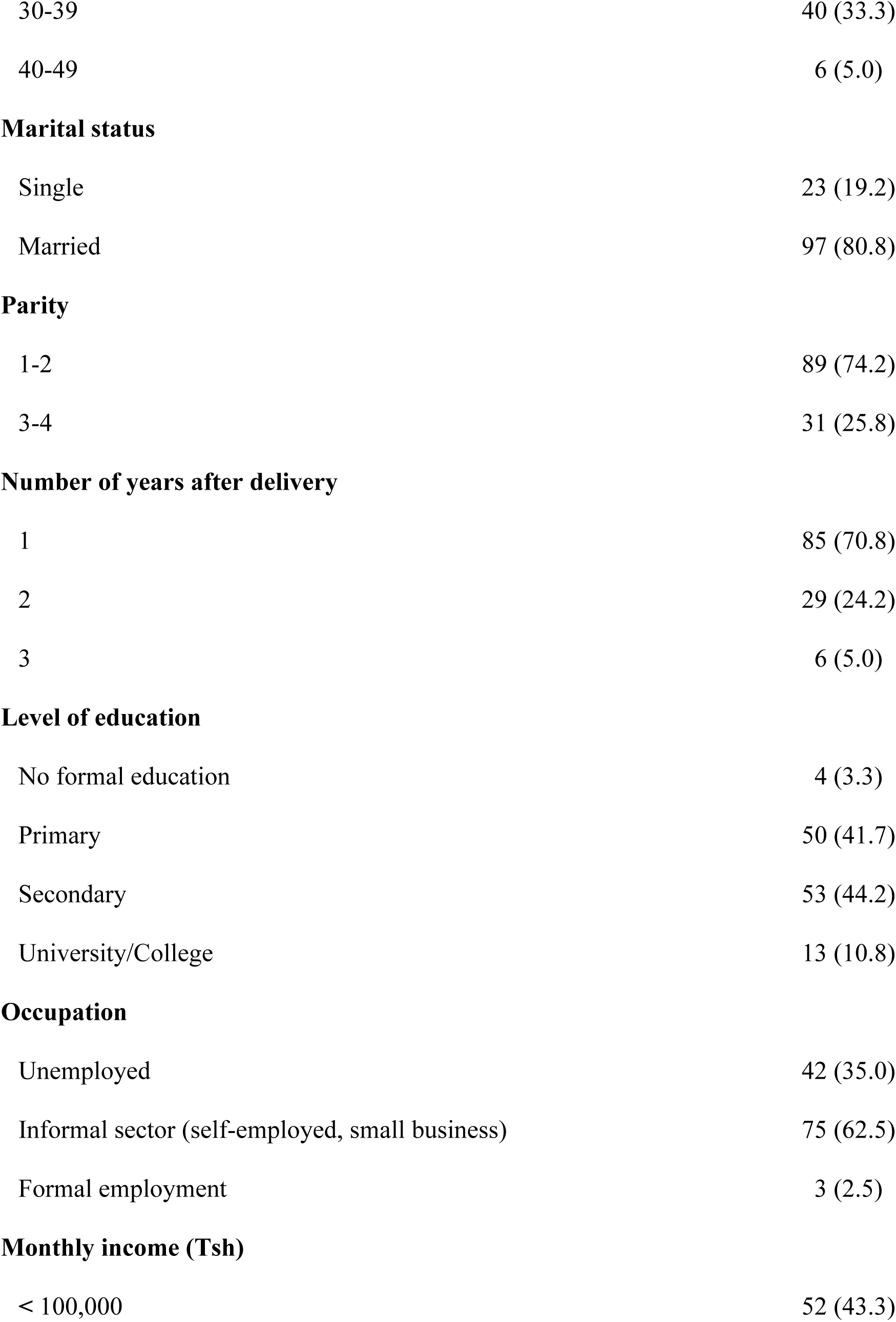
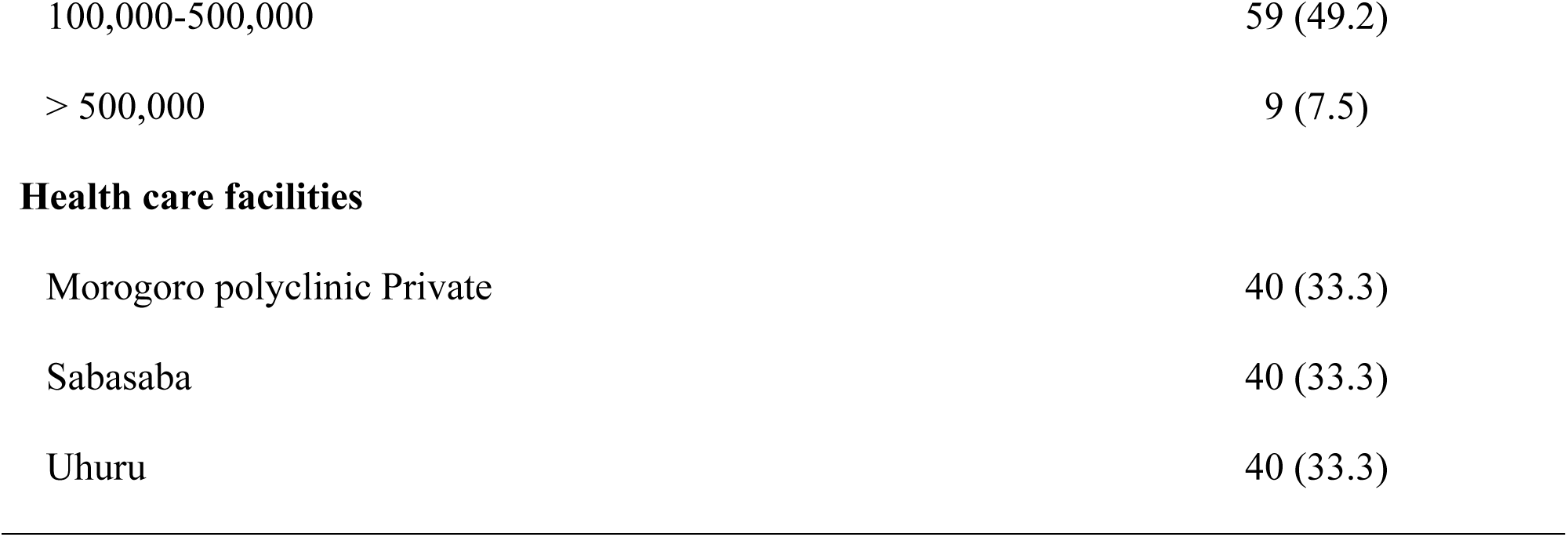
Socio-demographic characteristics of women beyond postpartum with children aged 1 to 5 years (n = 120).

| Characteristics | n (%) |
| --- | --- |
| **Age** |  |
| 15-19 | 8 (6.7) |
| 20-29 | 66 (55) |
| 30-39 | 40 (33.3) |
| 40-49 | 6 (5.0) |
| **Marital status** |  |
| Single | 23 (19.2) |
| Married | 97 (80.8) |
| **Parity** |  |
| 1-2 | 89 (74.2) |
| 3-4 | 31 (25.8) |
| **Number of years after delivery** |  |
| 1 | 85 (70.8) |
| 2 | 29 (24.2) |
| 3 | 6 (5.0) |
| **Level of education** |  |
| No formal education | 4 (3.3) |
| Primary | 50 (41.7) |
| Secondary | 53 (44.2) |
| University/College | 13 (10.8) |
| **Occupation** |  |
| Unemployed | 42 (35.0) |
| Informal sector (self-employed, small business) | 75 (62.5) |
| Formal employment | 3 (2.5) |
| **Monthly income (Tsh)** |  |
| < 100,000 | 52 (43.3) |
| 100,000-500,000 | 59 (49.2) |
| > 500,000 | 9 (7.5) |
| Health care facilities |  |
| Morogoro polyclinic Private | 40 (33.3) |
| Sabasaba | 40 (33.3) |
| Uhuru | 40 (33.3) |

### Prevalence of central obesity

The women beyond postpartum assessed had the mean waist circumference (WC), hip circumference (HC) and waist to hip ratio (WHR) of 87.2 ± 12.7 cm, 106.1 ± 13.1 cm and 0.82 ± 0.61 respectively. The prevalence of central obesity on the basis of waist circumference and waist to hip ratio were 68.3% and 37.5% respectively (Table 2).

**Table 2.** Prevalence of central obesity based on anthropometric measures (n = 120).

| Anthropometric measures | n (%) | Mean ± SD |
| --- | --- | --- |
| **Hip circumference** | | $106.1 \pm 13.1$ |
| **Waist circumference** | | $87.2 \pm 12.7$ |
| Centrally obese (>80) | 82 (68.3) |  |
| Not centrally obese (<80) | 38 (31.7) |  |
| **Waist to hip ratio** | | $0.82 \pm 0.61$ |
| Centrally obese | 45 (37.5) |  |
| Not centrally obese | 74 (61.7) |  |

**Table 3.**
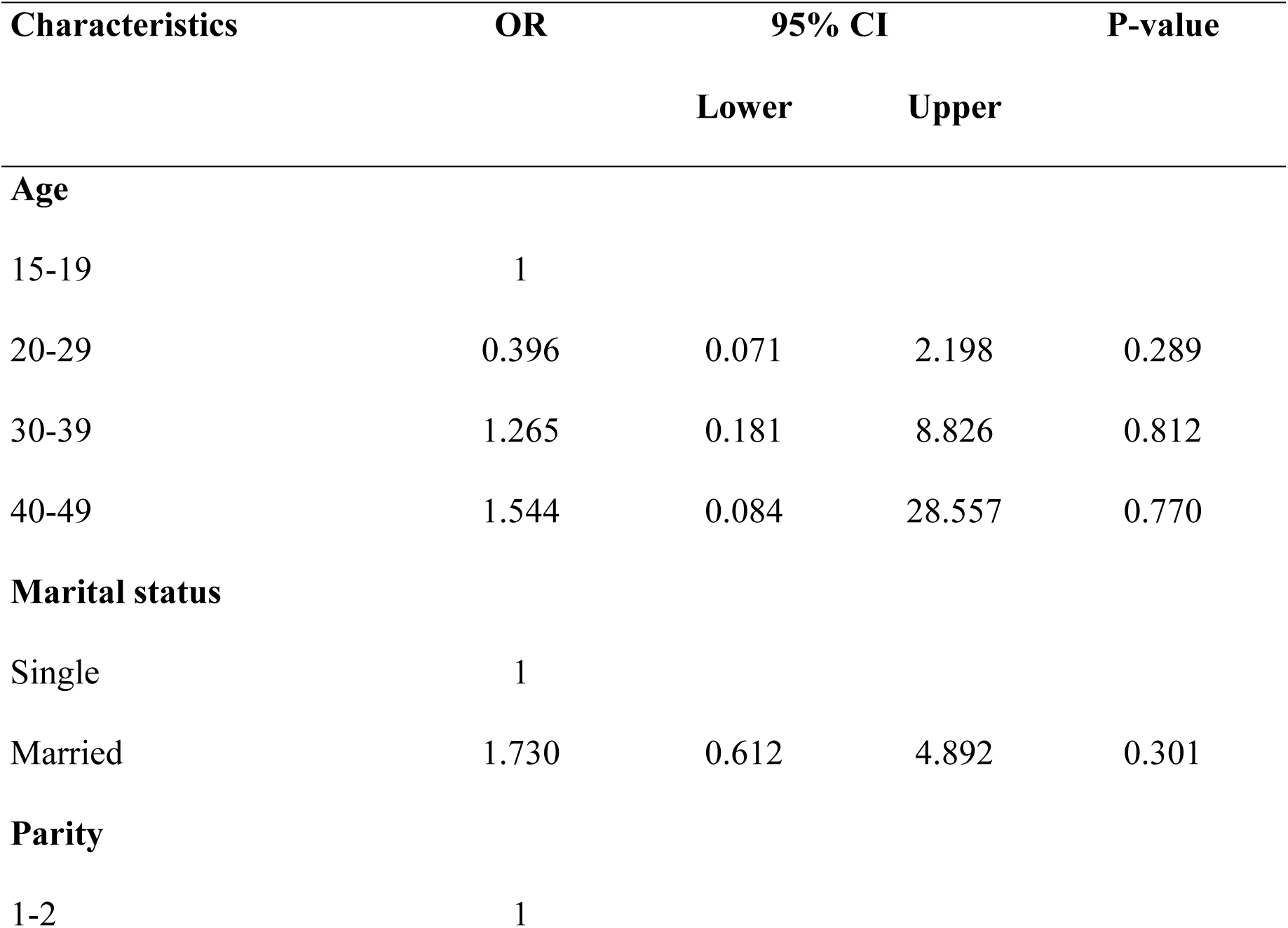
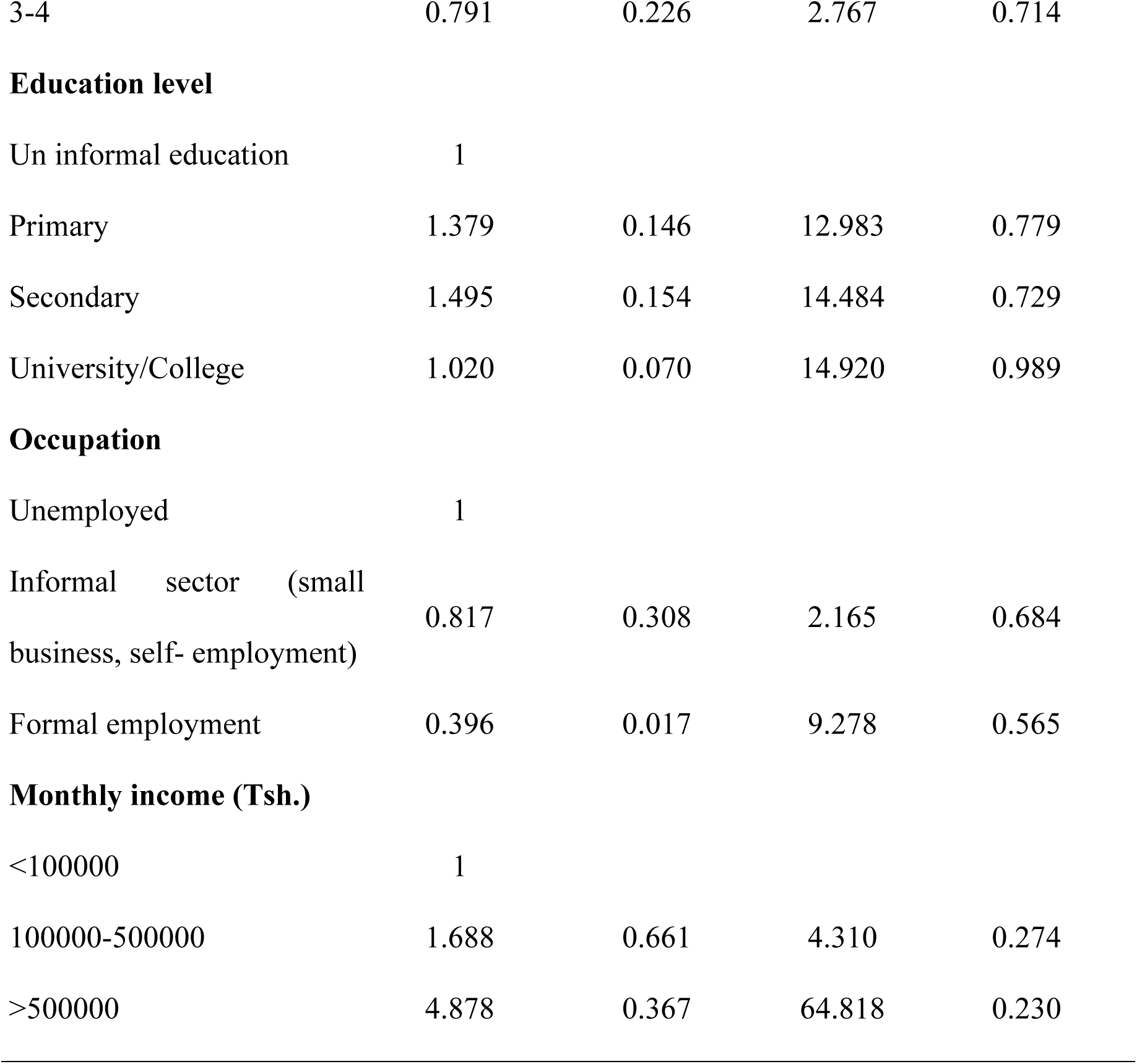
Relationship between central obesity and socio-demographic characteristics.

| Characteristics | OR | 95% CI |  | P-value |
| --- | --- | --- | --- | --- |
|  |  | Lower | Upper |  |
| Age |  |  |  |  |
| 15-19 | 1 |  |  |  |
| 20-29 | 0.396 | 0.071 | 2.198 | 0.289 |
| 30-39 | 1.265 | 0.181 | 8.826 | 0.812 |
| 40-49 | 1.544 | 0.084 | 28.557 | 0.770 |
| Marital status |  |  |  |  |
| Single | 1 |  |  |  |
| Married | 1.730 | 0.612 | 4.892 | 0.301 |
| Parity |  |  |  |  |
| 1-2 | 1 |  |  |  |
| 3-4 | 0.791 | 0.226 | 2.767 | 0.714 |
| **Education level** |  |  |  |  |
| Un informal education | 1 |  |  |  |
| Primary | 1.379 | 0.146 | 12.983 | 0.779 |
| Secondary | 1.495 | 0.154 | 14.484 | 0.729 |
| University/College | 1.020 | 0.070 | 14.920 | 0.989 |
| **Occupation** |  |  |  |  |
| Unemployed | 1 |  |  |  |
| Informal sector (small business, self- employment) | 0.817 | 0.308 | 2.165 | 0.684 |
| Formal employment | 0.396 | 0.017 | 9.278 | 0.565 |
| **Monthly income (Tsh.)** |  |  |  |  |
| <100000 | 1 |  |  |  |
| 100000-500000 | 1.688 | 0.661 | 4.310 | 0.274 |
| >500000 | 4.878 | 0.367 | 64.818 | 0.230 |

### Factors influencing central obesity

#### Relationship between central obesity and sociodemographic factors

None of the examined factors showed a statistically significant association with central obesity (p>0.05). However, participants aged 30–49 had higher odds of central obesity compared to those aged 20–29 (OR=1.544; 95% CI, 0.084-28.557).Married individuals had 1.73 times higher odds of central obesity than single individuals (OR = 0.788; 95% CI, 0.227-2.728), Those earning above TZS 500,000 had higher odds of having central obesity compared to those earning below TZS 100,000, though not statistically significant (OR=7.128; 95% CI, 0.571-89.058).

#### Lifestyle and dietary factors influencing central obesity

Nearly half of the subjects (42.5%) reported not consuming processed foods, while an equal proportion consumed fried foods one to two times per week. About 52.5% of the subjects consumed vegetables daily, while 65% consumed fruits one to two times per week.43.4% consume sugary drinks 1 to 2 times a week, 86.7% never engaged in physical activities and 30% had stress often. Almost (90%) of subjects did not pay attention to the portion sizes when having their meals, (Table 4).

**Table 4.**
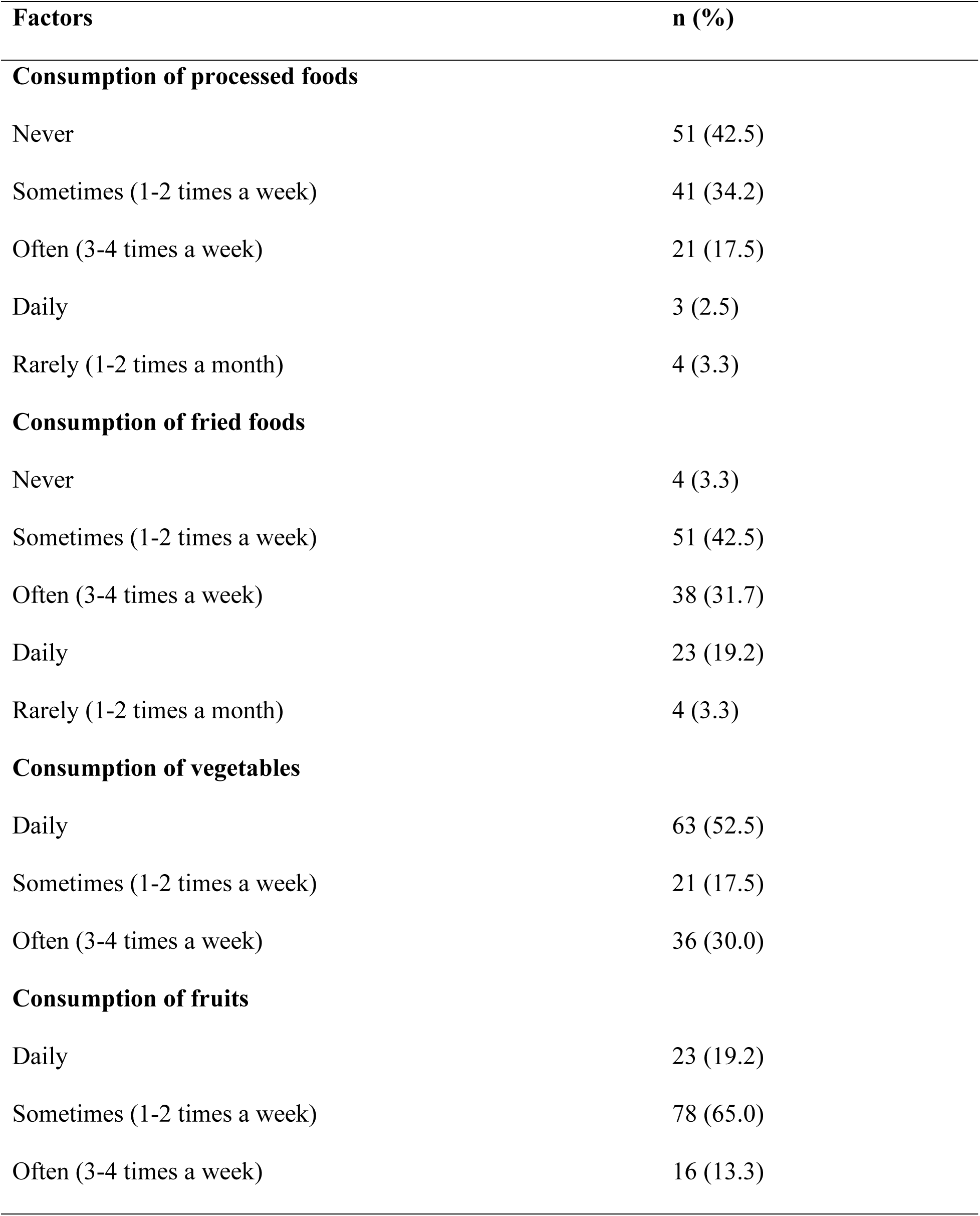
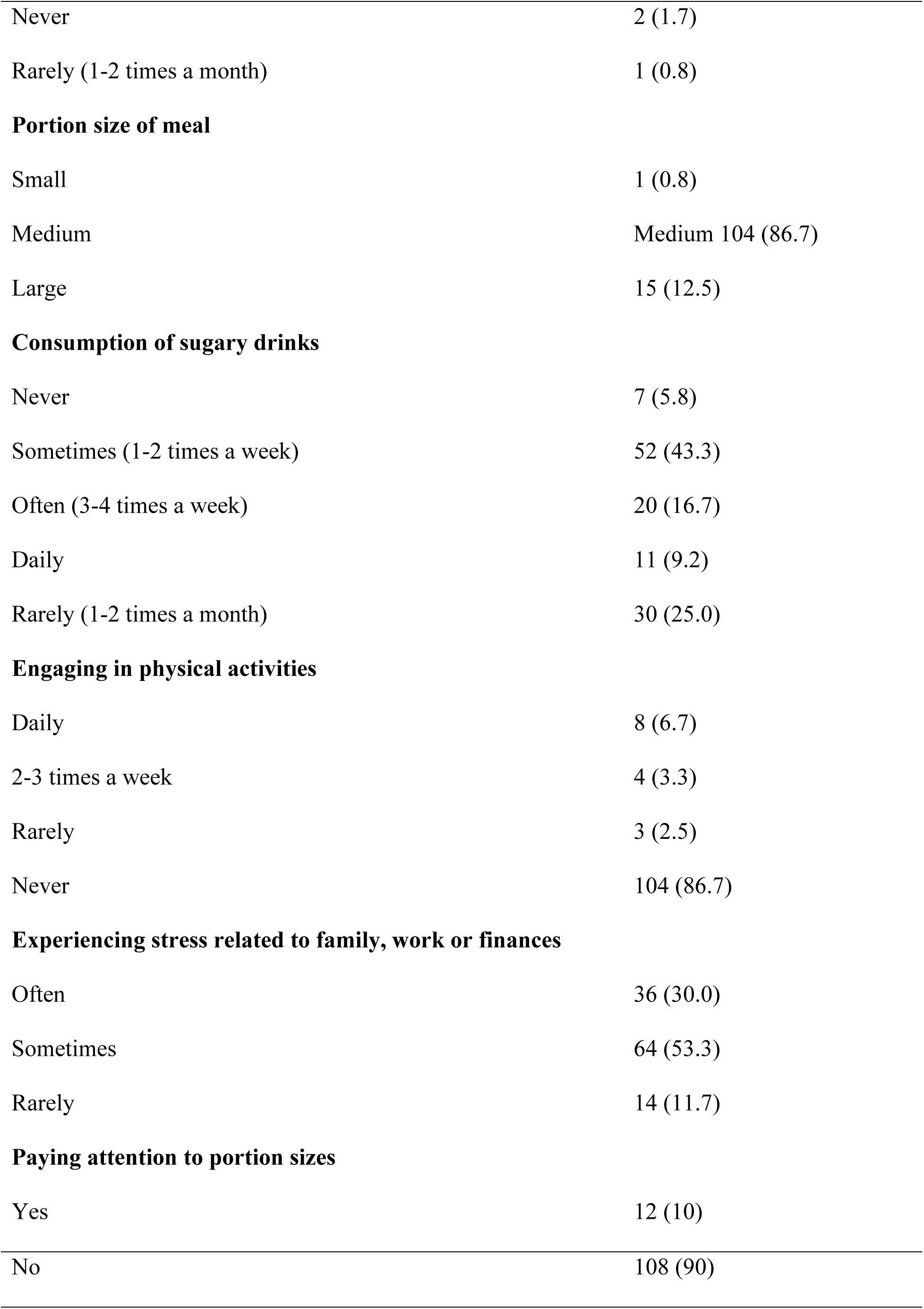
Lifestyle factors of the women beyond postpartum with children aged 1 to 5 years (n = 120).

| **Factors** | **n (%)** |
| --- | --- |
| **Consumption of processed foods** |  |
| Never | 51 (42.5) |
| Sometimes (1-2 times a week) | 41 (34.2) |
| Often (3-4 times a week) | 21 (17.5) |
| Daily | 3 (2.5) |
| Rarely (1-2 times a month) | 4 (3.3) |
| **Consumption of fried foods** |  |
| Never | 4 (3.3) |
| Sometimes (1-2 times a week) | 51 (42.5) |
| Often (3-4 times a week) | 38 (31.7) |
| Daily | 23 (19.2) |
| Rarely (1-2 times a month) | 4 (3.3) |
| **Consumption of vegetables** |  |
| Daily | 63 (52.5) |
| Sometimes (1-2 times a week) | 21 (17.5) |
| Often (3-4 times a week) | 36 (30.0) |
| **Consumption of fruits** |  |
| Daily | 23 (19.2) |
| Sometimes (1-2 times a week) | 78 (65.0) |
| Often (3-4 times a week) | 16 (13.3) |
| Never | 2 (1.7) |
| Rarely (1-2 times a month) | 1 (0.8) |
| **Portion size of meal** |  |
| Small | 1 (0.8) |
| Medium | Medium 104 (86.7) |
| Large | 15 (12.5) |
| **Consumption of sugary drinks** |  |
| Never | 7 (5.8) |
| Sometimes (1-2 times a week) | 52 (43.3) |
| Often (3-4 times a week) | 20 (16.7) |
| Daily | 11 (9.2) |
| Rarely (1-2 times a month) | 30 (25.0) |
| **Engaging in physical activities** |  |
| Daily | 8 (6.7) |
| 2-3 times a week | 4 (3.3) |
| Rarely | 3 (2.5) |
| Never | 104 (86.7) |
| **Experiencing stress related to family, work or finances** |  |
| Often | 36 (30.0) |
| Sometimes | 64 (53.3) |
| Rarely | 14 (11.7) |
| **Paying attention to portion sizes** |  |
| Yes | 12 (10) |
| No | 108 (90) |

### Increase in weight, waist size and challenges in managing central obesity

Most women beyond postpartum reported poor dietary choices (36.7%) and lack of time to exercise (25%) to be the reasons for the increase in weight and waist size (Fig 1). More than half (57.5%), of studied subjects reported to have lack of information on waist and weight management as the challenge in managing central obesity (Fig 2).

**Fig 1.**
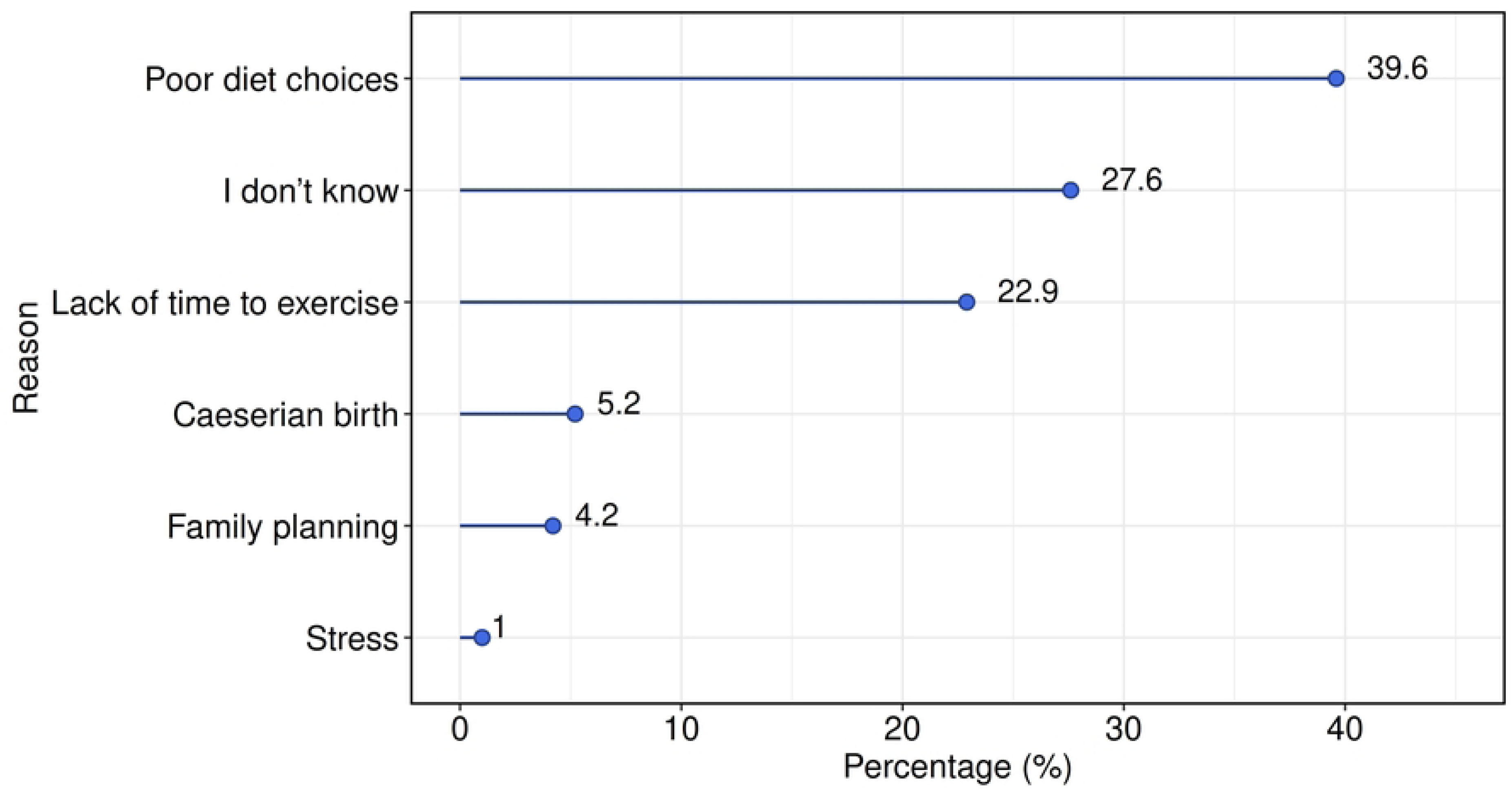
Reasons for the increase in weight and waist size as stated by past partum women.

**Fig 2.**
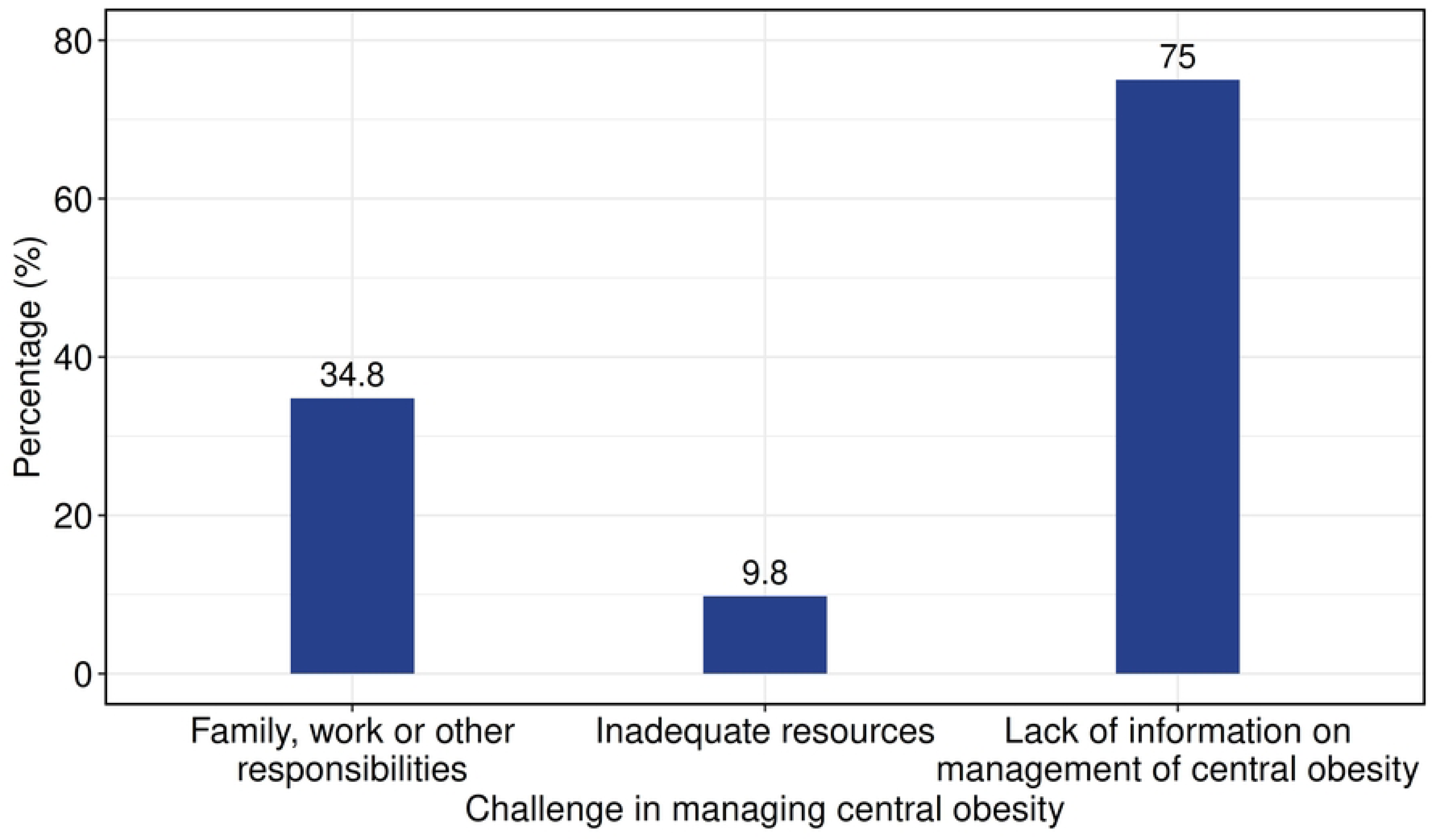
Challenges in managing central obesity by women beyond postpartum.

### Support from the health care systems

Nearly all of the studied subjects 94.2% and 95.0% respectively had not received any guidance from healthcare providers, nor support in managing or preventing central obesity, including nutrition counseling, exercise recommendations or monitoring of waist size and weight (Table 5).

**Table 5.**
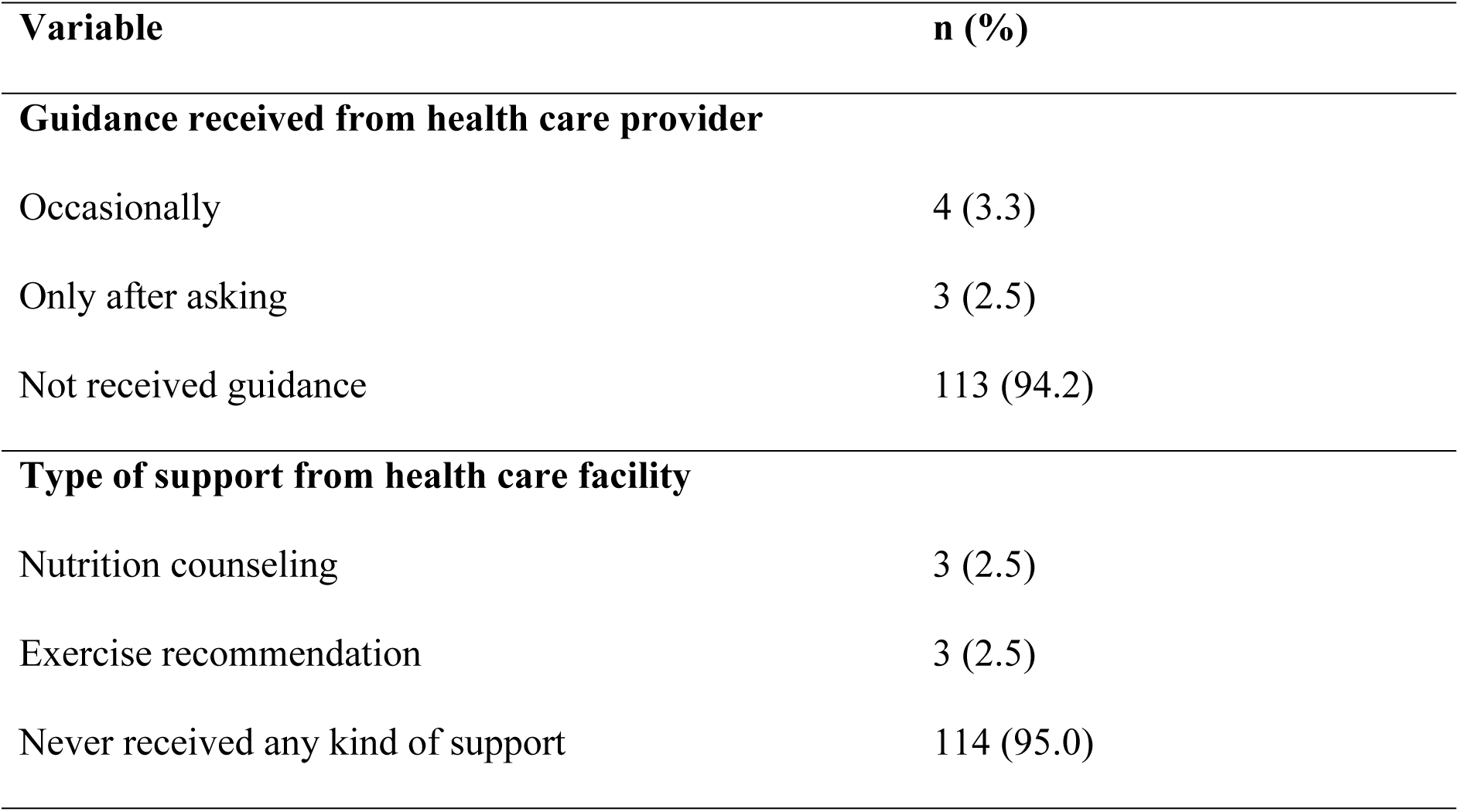
Guidance received from health care providers and type of support received from the facilities.

**Guidance received from health care provider**
|  |  |
| --- | --- |
| Occasionally | 4 (3.3) |
| Only after asking | 3 (2.5) |
| Not received guidance | 113 (94.2) |
| **Type of support from health care facility** |  |
| Nutrition counseling | 3 (2.5) |
| Exercise recommendation | 3 (2.5) |
| Never received any kind of support | 114 (95.0) |

## Discussion

### Central Obesity and Lifestyle Dietary Factors

This study showed a high prevalence of central obesity among women beyond postpartum. This is similar to other studies among women, where the prevalence of central obesity as defined by waist circumference was greater than fifty percent, where a study conducted by (28) prevalence of CO was 83.3%, 66.3% a study by (29), 51.7% a study by (30), 67.5% a study by (31) and 57.7% a study by (32). This could be contributed by sedentary lifestyles, poor dietary habits as many women have been observed to insufficient physical activity, intake of high fat and sugary foods and sleep deprivation that results to imbalance of ghrelin and leptin hormones thus stronger cravings for higher caloric foods (33).

Although none of the socio demographic factors showed a statistically significant association with central obesity, certain interesting patterns were noted. Older women were more likely to have central obesity where the possible reason for this could be reduced basal metabolism and physical activities as a result of ageing(34–36). In line with these findings, a study conducted in Southeast Ethiopia (37), Dodoma city (38), Kenya stepwise survey (39) and China (40) reported that the likelihood of having central obesity increased significantly with age. Also, it was observed that married women had higher chances of having central obesity compared to those who were not married which could be due to possibly changes in lifestyle and social roles after marriage (41). After marriage majority of the women have changes in their lifestyle including eating habits and social support as when they enter into marriage they often experience change in their daily routines that impact their health behaviors (42).

An increase in income was linked to higher odds of central obesity, showing that better financial status may lead to poor lifestyle behaviors such as consumption of energy dense foods, sugary drinks and reduced physical activity that may contribute to central obesity (43,44). A large proportion of women did not pay attention and monitor portion sizes of the meals when eating and therefore this has been associated with an increase in total energy intake thus greater opportunities to develop obesity (45). Very few studied subjects engaged in daily physical activities due to family and work responsibilities such as child caring, house chores and others being tight with income generation activities. Others reported not to engage in physical activity in belief that the responsibilities they have such as washing clothes and utensils, mopping, sweeping and going to the market in purchase of various stuffs was considered as practicing physical activity. These habits align with the energy imbalance theory, where energy intake exceeds energy expenditure, leading to fat accumulation around the abdomen (46).

### Challenges towards management of central obesity among past post-partum women

Participants identified poor dietary choices and lack of time to exercise as the main contributors to increase in weight and waist size. These poor dietary choices was highly related with cultural practices as they reported that after child delivery they commonly go to their in-laws where they are forced to eat a lot of food that is often fatty foods in belief of healing and having enough milk for breastfeeding their babies (24). Lack of time to exercise was also related with child caring and maternal responsibilities similar to the study conducted by (28). Also, women reported family work and responsibilities as among the challenges in managing weight and waist size. Many women especially in urban areas engage in income generation activities and they are also responsible for most of the house chores making them have a very tight schedule (47). This makes them have limited time to practice physical activity. The key informants in this study reported similar observation that women do not engage in physical exercise after child birth and they eat a lot of food in belief of getting enough milk to breastfeed their babies. Similarly, other studies have reported that women face time constraints due to child care responsibilities and prioritizing maternal responsibilities thus leading to limited time for meal planning and physical activity (48–50). However, many reported a lack of information on weight and waist management pointing to a critical gap in health education.

### Role of health care systems

Despite attending health care facilities, majority of the women had never received any guidance on weight or central obesity management or prevention. This was explained by the key informants to be caused by limited working staff in health care facilities that makes it hard for them to attend both children at Reproductive and Child Health (RCH) and women at once due to being overwhelmed with many duties. This aligns with the study by (51), where most women reported receiving no weight loss and no physical activity advice from a healthcare provider during the 3-month postpartum period. This serves as a challenge in support systems as the absence of structured follow up programs to support women in managing their weight post-delivery highlights the need for integrating central obesity prevention strategies into routine maternal and child health care services. Key informants also highlighted caesarean delivery as a significant barrier to weight and central obesity management among women beyond postpartum, giving factors such as prolonged recovery time, limited mobility and delayed return to physical activity all of which hinder efforts to engage in regular exercise and adopt healthy lifestyle practices. This was contrary to the study by (52) where they report that, caesarean delivery does not contribute to maternal weight retention instead driving factors such as pre-pregnancy weight and gestational weight gain and so recommending reversing the pattern of maternal weight gain through counselling and surveillance on weight management during child bearing years and between pregnancies in order to contribute to more stable weight trajectory for women throughout the life course. (53), reported that 12 months after delivery the women who had delivered by cesarean were more likely to have a postpartum weight retention of 10 pounds or more, than those who had a normal delivery.

### Limitations of this study

The facility-based sample that was used in the study may not be representative of all postpartum women, particularly those in rural areas or those who do not regularly access healthcare services hence failure to generalize results. Also, this study has not fully captured the cultural and societal influences on women’s health behaviors, which are important in understanding the broader context of past postpartum central obesity management.

## Conclusion

This study found a high prevalence of central obesity among women beyond postpartum, which is primarily influenced by unhealthy dietary practices, sedentary lifestyles and lack of support from health care systems. It was also observed that there is an increased risk of central obesity among older, married and higher-income in women beyond postpartum. The critical challenges that were faced by women beyond postpartum included; poor dietary habits, lack of time for physical activity, limited awareness on weight management and inadequate guidance from health care providers. Although participants had accessed health services, most had not been provided with nutrition counseling or recommendations on physical activity. Therefore, this study recommends the implementation of integrated postpartum and beyond postpartum care programs that include nutrition education, physical activity promotion and routine screening for central obesity. Strengthening the capacity of health care providers to deliver effective lifestyle counseling and support can play an important role in reducing the burden of central obesity among women beyond postpartum period. Also, nutrition education on central obesity management should be emphasized in community level so as to reach all women beyond postpartum.

## Data Availability

All relevant data are within the manuscript and its supporting information files

## Acknowledgements

The authors would like to thank the post-partum women, management and staff of the health facilities for their participation in the study. Lastly authors acknowledge everyone who contributed to the accomplishment of this work in one way or another.

## Author Contributions

Conceptualization: Rosela Remigius, Zalia O. Basheikh Data curation: Rosela Remigius, Rosula Remigius Formal analysis: Rosela Remigius

Methodology: Rosela Remigius, Zalia O. Basheikh Supervision: Zalia O. Basheikh

Validation: Rosela Remigius, Rosula Remigius, Zalia O. Basheikh Visualization: Rosela Remigius

Writing – original draft: Rosela Remigius

Writing – review & editing: Rosela Remigius, Rosula Remigius, Zalia O. Basheikh

## References

1. Peng W, Wang Y. Fighting obesity and non-communicable diseases needs different perspectives and new actions. Glob Health J. 2022 Sept 1;6(3):115–7.

2. Peng W, Chen S, Chen X, Ma Y, Wang T, Sun X, et al. Trends in major non-communicable diseases and related risk factors in China 2002–2019: an analysis of nationally representative survey data. Lancet Reg Health – West Pac [Internet]. 2024 Feb 1 [cited 2025 Nov 12];43. Available from: https://www.thelancet.com/journals/lanwpc/article/PIIS2666-6065(23)00127-X/fulltext

3. Wang Y, Xue H, Sun M, Zhu X, Zhao L, Yang Y. Prevention and control of obesity in China. Lancet Glob Health. 2019 Sept;7(9):e1166–7.

4. World Health Organization (WHO). Obesity [Internet]. 2025 [cited 2025 Nov 12]. Available from: https://www.who.int/health-topics/obesity

5. Hruby A, Hu FB. The Epidemiology of Obesity: A Big Picture. PharmacoEconomics. 2015 July;33(7):673–89.

6. Romieu I, Dossus L, Barquera S, Blottière HM, Franks PW, Gunter M, et al. Energy balance and obesity: what are the main drivers? Cancer Causes Control CCC. 2017 Mar;28(3):247–58.

7. Gesta S, Tseng YH, Kahn CR. Developmental origin of fat: tracking obesity to its source. Cell. 2007 Oct 19;131(2):242–56.

8. Ren H, Guo Y, Wang D, Kang X, Yuan G. Association of normal-weight central obesity with hypertension: a cross-sectional study from the China health and nutrition survey. BMC Cardiovasc Disord. 2023 Mar 8;23(1):120.

9. Cornier MA, Després JP, Davis N, Grossniklaus DA, Klein S, Lamarche B, et al. Assessing adiposity: a scientific statement from the American Heart Association. Circulation. 2011 Nov 1;124(18):1996–2019.

10. Rankinen T, Kim SY, Pérusse L, Després JP, Bouchard C. The prediction of abdominal visceral fat level from body composition and anthropometry: ROC analysis. Int J Obes Relat Metab Disord J Int Assoc Study Obes. 1999 Aug;23(8):801–9.

11. Mendoza MR, Flavia F, Hedley Q, Vel asquez IM. Prevalence of central obesity according to different definitions in normal weight adults of two cross-sectional studies in Panama. Lancet Reg Health - Am. 2022 June;10:100215.

12. Expert Panel on Detection, Evaluation, and Treatment of High Blood Cholesterol in Adults. Executive Summary of The Third Report of The National Cholesterol Education Program (NCEP) Expert Panel on Detection, Evaluation, And Treatment of High Blood Cholesterol In Adults (Adult Treatment Panel III). JAMA. 2001 May 16;285(19):2486–97.

13. Ezenwaka CE, Okoye O, Esonwune C, Onuoha P, Dioka C, Osuji C, et al. High prevalence of abdominal obesity increases the risk of the metabolic syndrome in Nigerian type 2 diabetes patients: using the International Diabetes Federation worldwide definition. Metab Syndr Relat Disord. 2014 June;12(5):277–82.

14. Zhang C, Rexrode KM, van Dam RM, Li TY, Hu FB. Abdominal obesity and the risk of all-cause, cardiovascular, and cancer mortality: sixteen years of follow-up in US women. Circulation. 2008 Apr 1;117(13):1658–67.

15. Katzmarzyk PT, Friedenreich C, Shiroma EJ, Lee IM. Physical inactivity and non-communicable disease burden in low-income, middle-income and high-income countries. Br J Sports Med. 2022 Jan;56(2):101–6.

16. Meyer HE, Willett WC, Flint AJ, Feskanich D. Abdominal obesity and hip fracture: results from the Nurses’ Health Study and the Health Professionals Follow-up Study. Osteoporos Int J Establ Result Coop Eur Found Osteoporos Natl Osteoporos Found USA. 2016 June;27(6):2127–36.

17. Wong MCS, Huang J, Wang J, Chan PSF, Lok V, Chen X, et al. Global, regional and time-trend prevalence of central obesity: a systematic review and meta-analysis of 13.2 million subjects. Eur J Epidemiol. 2020;35(7):673–83.

18. Kabwama SN, Kirunda B, Mutungi G, Wesonga R, Bahendeka SK, Guwatudde D. Prevalence and correlates of abdominal obesity among adults in Uganda: findings from a national cross-sectional, population based survey 2014. BMC Obes. 2018;5:40.

19. Olatunbosun ST, Kaufman JS, Bella AF. Central Obesity in Africans: Anthropometric Assessment of Abdominal Adiposity and its Predictors in Urban Nigerians. J Natl Med Assoc. 2018 Oct;110(5):519–27.

20. Malik SK, Kouame J, Gbane M, Coulibaly M, Ake MD, Ake O. Prevalence of abdominal obesity and its correlates among adults in a peri-urban population of West Africa. AIMS Public Health. 2019;6(3):334–44.

21. Owolabi EO, Ter Goon D, Adeniyi OV. Central obesity and normal-weight central obesity among adults attending healthcare facilities in Buffalo City Metropolitan Municipality, South Africa: a cross-sectional study. J Health Popul Nutr. 2017 Dec 28;36(1):54.

22. Munyogwa MJ, Ntalima KS, Kapalata SN. Setting – based prevalence and correlates of central obesity: findings from a cross-sectional study among formal sector employees in Dodoma City, Central Tanzania. BMC Public Health. 2021 Jan 7;21:97.

23. McKinley MC, Allen-Walker V, McGirr C, Rooney C, Woodside JV. Weight loss after pregnancy: challenges and opportunities. Nutr Res Rev. 2018 Dec;31(2):225–38.

24. Jaakkola J, Hakala P, Isolauri E, Poussa T, Laitinen K. Eating behavior influences diet, weight, and central obesity in women after pregnancy. Nutrition. 2013 Oct;29(10):1209–13.

25. Scott C, Andersen CT, Valdez N, Mardones F, Nohr EA, Poston L, et al. No global consensus: a cross-sectional survey of maternal weight policies. BMC Pregnancy Childbirth. 2014 May 15;14(1):167.

26. Makama M, Skouteris H, Moran LJ, Lim S. Reducing Postpartum Weight Retention: A Review of the Implementation Challenges of Postpartum Lifestyle Interventions. J Clin Med. 2021 Apr 27;10(9):1891.

27. Morogoro Urban District. Morogoro Urban District. In: Wikipedia [Internet]. 2024 [cited 2025 July 24]. Available from: https://en.wikipedia.org/w/index.php?title=Morogoro_Urban_District&oldid=1254570027

28. Manikandan N, Vasudevan V, Elayaperumal S. PREVALENCE OF CENTRAL OBESITY AMONG POST PARTUM WOMEN – A CROSS SECTIONAL OBSERVATIONAL STUDY. Obstet Gynaecol Forum. 2024 May 13;34(2s):64–7.

29. Cherono R, Ogada IA, Kimiywe J. Weight Status at Postpartum: Being Normal Weight Yet Centrally Obese! Food Nutr Sci. 2019 Sept 3;10(9):1085–95.

30. Du P, Wang HJ, Zhang B, Qi SF, Mi YJ, Liu DW, et al. Prevalence of abdominal obesity among Chinese adults in 2011. J Epidemiol. 2017 June 1;27(6):282–6.

31. Mbochi RW, Kuria E, Kimiywe J, Ochola S, Steyn NP. Predictors of overweight and obesity in adult women in Nairobi Province, Kenya. [Internet]. 2012 [cited 2025 July 4]. Available from: http://archive.org/details/pubmed-PMC3485189

32. Damorou F, Yayehd K, N’cho Mott MP, Tcherou T, Ehlan E, N’da NW, et al. Prevalence and Determinants of Obesity among Workers in Lomé (Togo). Res J Cardiol. 2012 Dec 15;6(1):19–27.

33. Capistrano D, Sison G. 11 Best Online TRT Clinics in 2025: Don’t Get Tricked [Internet]. 2025 [cited 2025 Oct 7]. Available from: https://www.nutritionnc.com/best-online-trt-clinics/

34. Biru B, Taye A, Feyisa RB. Central obesity and its predictors among adults in Nekemte town, West Ethiopia [Internet]. 2021 [cited 2025 Oct 8]. Available from: https://scholar.google.com/scholar_lookup?author=B+Biru&author=D+Tamiru&author=A+Taye&author=B.+Regassa+Feyisa&title=Central+obesity+and+its+predictors+among+adults+in+Nekemte+town%2C+West+Ethiopia&publication_year=2021&journal=SAGE+open+medicine&volume=9

35. Henry CJ. Mechanisms of changes in basal metabolism during ageing. Eur J Clin Nutr. 2000 June;54 Suppl 3:S77–91.

36. Malik SK, Kouame J, Gbane M, Coulibaly M, Ake MD, Ake O, et al. Prevalence of abdominal obesity and its correlates among adults in a peri-urban population of West Africa. AIMS Public Health. 2019;6(3):334–44.

37. Tekalegn Y, Solomon D, Sahiledengle B, Assefa T, Negash W, Tahir A, et al. Prevalence of central obesity and its associated risk factors among adults in Southeast Ethiopia: A community-based cross-sectional study. PLOS ONE. 2022 Aug 5;17(8):e0265107.

38. Munyogwa MJ, Ntalima KS, Kapalata SN. Setting – based prevalence and correlates of central obesity: findings from a cross-sectional study among formal sector employees in Dodoma City, Central Tanzania. BMC Public Health. 2021 Jan 7;21:97.

39. Nyakundi C, Okemwa S, Ngesa RW, Gatimu SM. Prevalence and determinants of central obesity among adults 18–69 years in Kenya: a cross-sectional study [Internet]. medRxiv; 2024 [cited 2025 July 4]. p. 2024.09.18.24313881. Available from: https://www.medrxiv.org/content/10.1101/2024.09.18.24313881v1

40. Nan J, Chen M, Yuan H, Cai S, Piao W, Li F, et al. Prevalence and Influencing Factors of Central Obesity among Adults in China: China Nutrition and Health Surveillance (2015–2017). Nutrients. 2024 Jan;16(16):2623.

41. Omar SM, Taha Z, Hassan AA, Al-Wutayd O, Adam I. Prevalence and factors associated with overweight and central obesity among adults in the Eastern Sudan. PLoS ONE. 2020 Apr 30;15(4):e0232624.

42. Nikolic Turnic T, Jakovljevic V, Strizhkova Z, Polukhin N, Ryaboy D, Kartashova M, et al. The Association between Marital Status and Obesity: A Systematic Review and Meta-Analysis. Diseases. 2024 July 5;12(7):146.

43. Martey E, Aheto JMK, Afesorgbor SK. The Conversation. 2024 [cited 2025 July 19]. Economic development in sub-Saharan Africa is linked to increasing obesity rates in women. Available from: http://theconversation.com/economic-development-in-sub-saharan-africa-is-linked-to-increasing-obesity-rates-in-women-230745

44. Mosha D, Paulo HA, Mwanyika-Sando M, Mboya IB, Madzorera I, Leyna GH, et al. Risk factors for overweight and obesity among women of reproductive age in Dar es Salaam, Tanzania | BMC Nutrition [Internet]. 2021 [cited 2025 July 19]. Available from: https://link.springer.com/article/10.1186/s40795-021-00445-z?utm_source=chatgpt.com

45. Sievert K, Hussain SM, Page MJ, Wang Y, Hughes HJ, Malek M, et al. Effect of breakfast on weight and energy intake: systematic review and meta-analysis of randomised controlled trials. BMJ. 2019 Jan 30;364:l42.

46. Cabbage S, Tewari S. Chapter 9: Energy Balance. 2025 [cited 2025 July 5]; Available from: https://spscc.pressbooks.pub/principlesofnutrition/part/chapter-9-energy-balance/

47. Ngai RL, Dinkelman T. Female time use and structural transformation in Africa | CEPR [Internet]. 2022 [cited 2025 July 19]. Available from: https://cepr.org/voxeu/columns/female-time-use-and-structural-transformation-africa?utm_source=chatgpt.com

48. Lim S, Lang S, Savaglio M, Skouteris H, Moran LJ. Intervention Strategies to Address Barriers and Facilitators to a Healthy Lifestyle Using the Behaviour Change Wheel: A Qualitative Analysis of the Perspectives of Postpartum Women. Nutrients. 2024 Jan;16(7):1046.

49. Makama M, Skouteris H, Moran LJ, Lim S. Reducing Postpartum Weight Retention: A Review of the Implementation Challenges of Postpartum Lifestyle Interventions. J Clin Med. 2021 Apr 27;10(9):1891.

50. Ryan RA, Lappen H, Bihuniak JD. Barriers and Facilitators to Healthy Eating and Physical Activity Postpartum: A Qualitative Systematic Review. J Acad Nutr Diet. 2022 Mar;122(3):602–613.e2.

51. Ferrari RM, Siega-Riz AM, Evenson KR, Moos MK, Melvin CL, Herring AH. Provider Advice About Weight Loss and Physical Activity in the Postpartum Period. J Womens Health. 2010 Mar;19(3):397–406.

52. Kapinos A K, Yakusheva O, Weiss M. Cesarean deliveries and maternal weight retention | BMC Pregnancy and Childbirth | Full Text [Internet]. 2017 [cited 2025 Oct 8]. Available from: https://bmcpregnancychildbirth.biomedcentral.com/articles/10.1186/s12884-017-1527-x

53. Legro NR, Lehman EB, Kjerulff KH. Mode of first delivery and postpartum weight retention at 1 year. Obes Res Clin Pract. 2020;14(3):241–8.

